# Associations Between Daily Outdoor Temperature and Subjective Real-time Ratings of Emotional States and Sleep in Mood Disorder Subtypes

**DOI:** 10.1101/2025.09.08.25335358

**Authors:** Debangan Dey, Hamza A. Lateef, Andrew Leroux, Vadim Zipunnikov, Kathleen Merikangas

## Abstract

Growing evidence for the influence of weather on mental health at both the aggregate level indices of mental health statistics of hospitalizations, morbidity, and mortality, and individual level dynamics of mood states. Most research on this topic has focused on light exposure and depressed mood as the sole indicators of seasonal fluctuations of mood disorders. This paper evaluates the association between daily maximum outdoor temperature (DMOT) and contemporaneously reported mood, energy, anxiousness, and sleep quality assessed with Ecological Momentary Assessment (EMA) in a community-based sample of 452 people, aged 11 to 85 years, comprising people with or without a history of mood disorders. After controlling for demographics, daily activity levels, and daytime cloud coverage as an index of light exposure, we found that higher DMOT was associated with better mood, increased energy, and better sleep quality among those with with a history of mood disorders, particularly in the spring among people with a history of Bipolar Disorder (BD) who tended to exhibit more seasonal changes than controls. These findings suggest that the dynamics of mood, energy, and sleep may underlie the aggregate population-level influences of temperature and correlated environmental influences on negative mental health outcomes, particularly among those with BD. Therefore, the conceptualization of risk and intervention for mood disorders should consider both the aggregate influences of temperature and light and associated environmental conditions, as well as individual-level sleep quality and energy as potential mechanisms for seasonal patterns of emotional states.

**HIGHLIGHTS:**

- Higher spring temperatures improved mood, energy, and sleep quality in BD and MDD.
- Fall temperatures showed U-shaped associations with mood and energy in BD.
- Temperature effects varied by mood disorder subtype and season.
- Findings support incorporating environmental factors in mood disorder management.

## INTRODUCTION

There have been widespread efforts to investigate the influences of patterns of weather and climate, as well as acute exposures to weather-related events and environmental toxins on human physiology and disease (Campbell-Lendrum et al., 2023). In particular, weather has been shown to have a major impact on mental health in terms of both aggregate evidence for hospitalizations, psychiatric emergency room visits, and suicides, and individual-level perception of well-being and mood states (Radua et al., 2024). However, attributions regarding potential mechanisms are complicated by the complexity of the key factors involved in climate, including temperature, light, weather, air and water pollution, and their inter-relationships.

Light exposure has been one of the most widely studied environmental factors associated with both physical and mental health. In a study of more than 85,000 adults across the world, exposure to artificial light at night (ALAN) was associated with several mental disorders (Burns et al., 2023), and with poorer sleep, anxiousness, and depression, particularly among youth with bipolar disorder in a nationally representative sample of adolescents in the US (Paksarian et al., 2020). The widespread recognition of seasonal patterns of depression, with an apparent peak in winter among individuals with mood disorders, has even been embedded as a sub-category of mood disorder in the contemporary diagnostic nomenclature (DSM-5). However, this purported winter peak has not been evident in aggregate statistics from multiple sources that exhibit the peak incidence of suicides and hospitalizations for depression in the spring, with a secondary peak in the fall (To et al., 2024; Yu et al., 2020). This discrepancy between individual-level and aggregate-level findings suggests that the underlying explanations for seasonal patterns may be more complex than light exposure alone. In particular, the influence of ambient temperature, which is highly correlated with seasonal light patterns, has rarely been considered in studies of seasonal mood disorders.

There is compelling evidence for an association between ambient temperature and several mental health-related events. Numerous systematic reviews of evidence regarding climate and temperature (Thompson et al., 2018); (Clery et al., 2024; Liu et al., 2021; Radua et al., 2024; Thompson et al., 2023) have concluded that ambient temperature is associated with numerous population-level indicators of mental health outcomes, including hospitalizations, emergency room visits, and completed and attempted suicides. Likewise, qualitative analyses of individual-level data have yielded significant associations between higher temperature and diverse subjective mental health outcomes, including psychological distress, negative well-being, negative affect, and depressive symptoms, albeit with substantial heterogeneity in estimated effects with respect to the measurement of temperature and mental health, as well as world region and latitude and demographic factors, particular age (Liu et al., 2021).

Studies of the individual-level impact of light and temperature consistently show that the susceptibility to environmental perturbations may differ among those with mental disorders. In particular, people with bipolar disorder have increased sensitivity to both light (Roguski et al., 2024) and temperature (Montes et al., 2021). However, there is still a lack of prospective data from controlled studies on the influences of diverse environmental factors on mood states in people with mood disorders. Recent secondary analyses of prospective daily recorded mood suggested that increasing temperature was associated with increased sad mood, particularly among people with bipolar disorder (Bundo et al., 2023). Similar analyses of short-term fluctuations in emotional states rated on a smartphone application in a large sample of people with either depression or bipolar disorder showed that higher ambient temperatures were associated with lower depressive symptom scores and higher mania symptom ratings (Clery et al., 2024). However, these analyses did not consider the potential confounding influence of daytime light exposure.

There are several limitations to the evidence regarding the associations between weather and mood states and disorders. First, the strongest evidence derived from aggregate-level associations reported in meta-analyses does not include potential confounders either at the aggregate or individual level.

Second, the majority of individual-level data have come from non-systematic samples obtained through either telephone surveys (Noelke et al., 2016; Obradovich et al., 2018) or social media, modalities which are both subject to sampling and recall biases (Baylis et al., 2018);(Ruths & Pfeffer, 2014). Third, most studies have examined broad outcomes such as psychological distress, negative affect, or negative well-being without consideration of clinically significant mood or other mental disorders. Fourth, individual level studies rarely considered potential correlates or mechanisms for mood changes such as sleep that may comprise a common mechanism for broad changes in mental health outcomes associated with temperature fluctuations (Chevance et al., 2024). Fifth, most studies focus on light or temperature alone without considering their potential joint influences nor other highly intercorrelated environmental factors such as season, noise, pollution, or humidity. Finally, there is substantial heterogeneity (differences in significance and signs) in these findings that may vary due to differences in sample characteristics such as age and climate zones (Liu et al., 2021;Thompson et al., 2018).

The current work addresses several key limitations of previous studies, including a well-characterized community-based sample for specific mood disorder subtypes, inclusion of comparison groups of people without mood disorders, prospective within and between-day tracking of mood and other emotional states, and application of aggregate exposure to both temperature and light. The specific aims of this paper are to: (1) examine the association between daily maximum outdoor temperature (DMOT) on four self-reported emotional states (sad mood, energy, anxiousness, and sleep quality) administered with Ecological Momentary Assessment (EMA) after adjustment for demographics, season (winter, spring, summer, fall), and daytime light; and (2) evaluate whether these associations differ by a lifetime history of mood disorder subtypes including Bipolar Disorder (BD) and Major Depressive Disorder.

## METHODS

### Sample

Our full sample consisted of 502 participants from the National Institute of Mental Health Family Study, aged 11 to 85 years. Recruitment and screening of participants occurred within the greater Washington DC metropolitan area community, involving outreach to 11,000 households within a 50-mile radius of Washington DC, as well as the NIH Clinical Center general volunteer referral core, local health newsletters and announcements, and individuals from the NIMH Mood and Anxiety Disorder Program. All participants provided written informed consent for the protocol approved by the Combined Neuroscience IRB at the National Institutes of Health (NIH; protocol no. 03-M-0211). Further details about the NIMH Family Study can be found in (Ballard et al., 2019; Lamers et al., 2018; Merikangas et al., 2014, 2019).

### Ecological Momentary Assessment (EMA)

EMA (Ecological Momentary Assessment) procedures were utilized to gather a comprehensive range of domains, encompassing the mood circumplex, context, life events, sleep, physical activity, eating/drinking, pain, and headache, rated on a research-administered Android smartphone. We included the analog ratings (1-7 scale) of sad mood, anxiousness, and energy assessed by the mood circumplex (Larsen & Diener, 1992). Participants were prompted to rate their current emotional states four times a day on a Likert scale ranging from (1) to (7) for anxiousness [(1) very calm to (7) very anxious], sad mood [(1) very happy to (7) very sad], and energy [(1) very tired to (7) very energetic]. Additionally, at the first assessment of the day, participants rated the quality of their sleep for the previous night [(1) not at all rested to (7) fully rested]. More details are provided in prior publications of EMA in this study (Lamers et al., 2018).

### Weather data

Daily maximum outdoor temperature (DMOT) was collected using gridded (∼ 4km) temperature data from the PRISM climate group (Di Luzio et al., 2008). Average daily total cloud cover (measured in percentages) data was collected from <u>NCEP North American Regional Reanalysis</u> <u>(NARR) website</u>. DMOT and cloud cover data at the median latitude-longitude of the zip code associated with participants’ residential addresses during the study were linked with participant EMA data.

### Seasons

Analyses were stratified by season due to the intrinsic covariation of season, light, and DMOT, avoiding issues of interpretations based on extrapolation (i.e., a DMOT of 10 degrees F in summer in the Washington DC region). Specifically, four seasons were defined. 1. Spring (15th Mar - 14th Jun), 2. Summer (15th Jun - 14th Sep), 3. Fall (15th Sep - 14th Dec), 4. Winter (15th Dec - 14th Mar). Given the potential differential effects of temperature on the mood circumplex by specific seasons or mood disorder subgroups, we conducted stratified analyses by seasons and mood disorders. In consideration of statistical power, mood disorder diagnosis groups were partially collapsed for all analyses. Participants with either Bipolar I or Bipolar II were grouped into a single Bipolar Disorder category. Additionally, participants with an Anxiety disorder but without a mood disorder diagnosis were grouped with unaffected controls. Participants with assessments across two seasons, we assigned the season in which they had the majority of assessments.

### Mental Disorders

Mood and Anxiety disorders were evaluated through an extensive diagnostic assessment utilizing the NIMH Family Study Diagnostic Interview for Affective Spectrum Disorders (DIAS) (Merikangas et al., 2014). The mood disorder subgroups were characterized by mutually exclusive mood disorder subtypes, including Bipolar I (BPI) (n=54), Bipolar II (BPII) (n=52), and Major Depressive Disorder (MDD)(n=143). The two groups with bipolar disorder were combined for these analyses. There were a total of 203 controls who had no history of mood disorders. For participants with assessments across two seasons, we assigned the season in which they had the majority of assessments.

### Statistical methods

Associations between EMA items (self-reported mood, anxiousness, energy, and sleep quality) and DMOT were estimated with the Generalized additive mixed model framework (Hastie & Tibshirani, 1986), implemented in the mgcv package (S. N. Wood, 2017; S. Wood & Wood, 2015) in the R statistical software 4.2. All models adjusted for daily total cloud cover to account for potential confounding of observed associations with exposure to outdoor daytime light, as well as age, sex, and lifetime mood disorder subgroup. Within-person correlation was accounted for statistically using subject-specific random intercepts. Non-linear associations between DMOT and EMA responses were modeled using rank 9 penalized B-splines with a second-order smoothness penalty. Differential associations by mood disorder groups were assessed using a varying coefficient model, again estimated using rank 9 penalized splines, allowing for potentially mood-disorder-specific non-linear effects. The models estimated are described in precise mathematical detail in the supplemental materials, Section 2.

Results are presented as percentage changes in each outcome (EMA response) relative to its average value at the median temperature for each season (Figures 1, 2). Specifically, we calculated the percentage change by comparing the model-predicted value of the outcome at a given temperature to the predicted value at the median temperature for that season. The median temperatures for each season were 5°C for winter, 20°C for spring, 30°C for summer, and 15°C for fall. This approach allows us to interpret how changes in temperature during a specific season contribute to the percentage change in mood, anxiousness, energy, or sleep quality. Estimates, p-values, and pointwise Wald 95% confidence intervals were obtained for estimated associations and plotted for all models. Statistical significance was assessed using a threshold of 0.05 against a null hypothesis of no significant effect.

**Fig 1:**
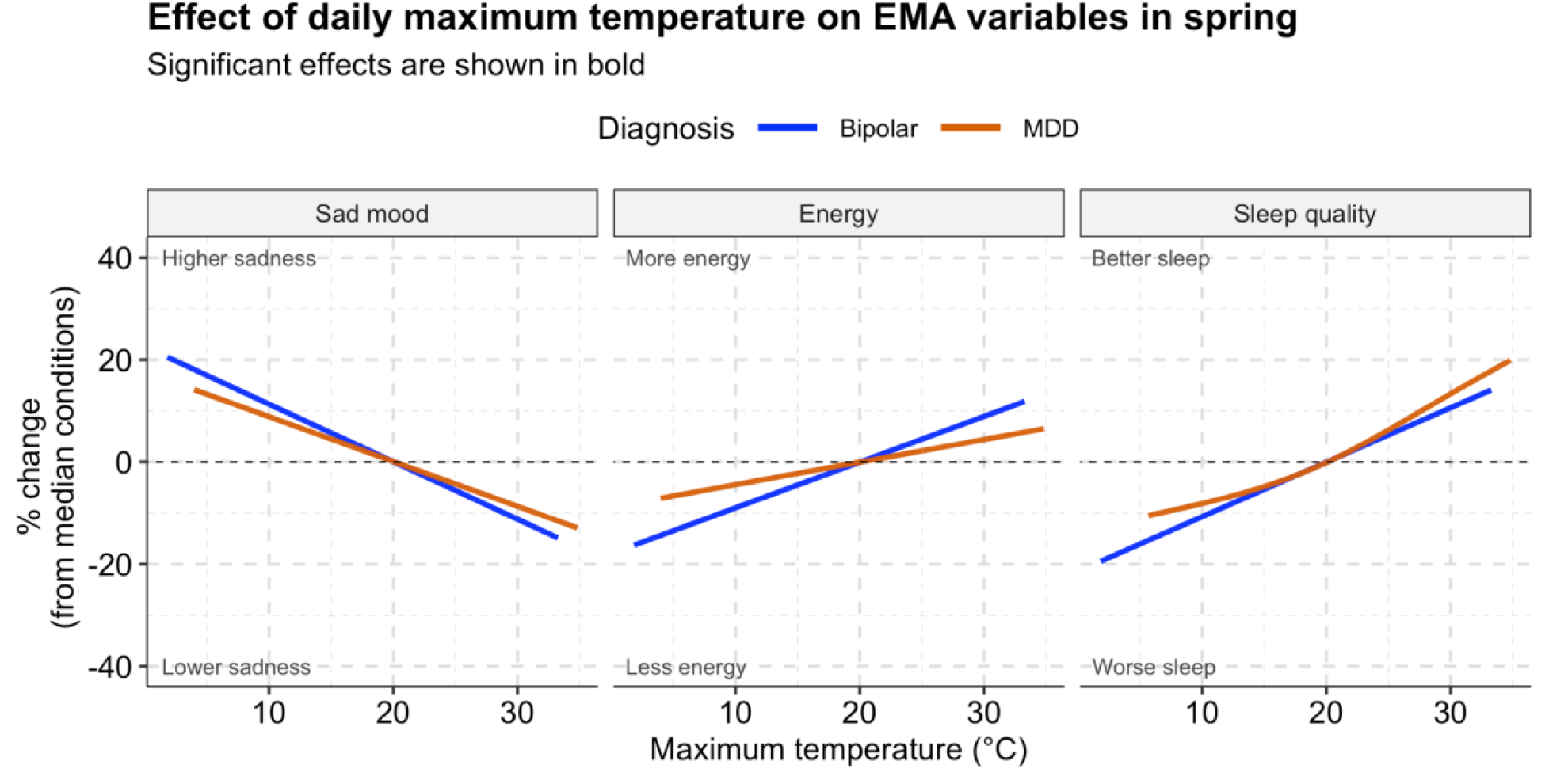
Spring: Significant associations of daily maximum outdoor temperature (DMOT) with self-reported sad mood (left), energy (middle), and sleep quality (right) among participants with Bipolar and MDD.

**Fig 2:**
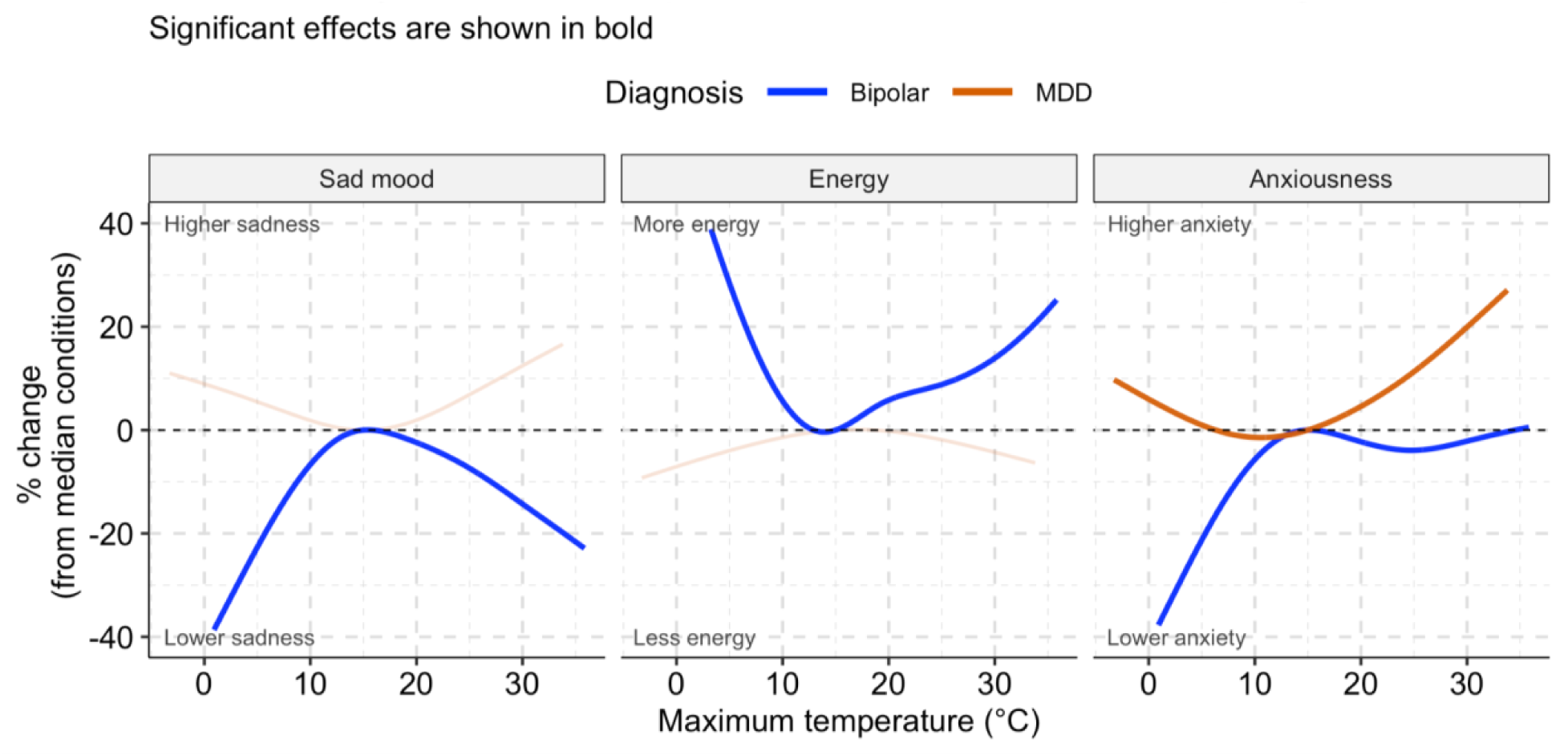
Fall: Significant associations of daily maximum outdoor temperature (DMOT) with self-reported sad mood (left), energy (middle), and anxiousness (right) among participants with a history of Bipolar and MDD.

## RESULTS

### Demographics and clinical history of participants

The full sample included 502 participants, contributing a total of 7,028 observations. We excluded 50 participants (9.7%) who lacked location information, or did not have at least two days of complete data on both EMA variables (sad mood, anxiousness, energy, and sleep quality) and corresponding temperature measurements. After applying our inclusion criteria, the analytic dataset contained 6328 daily assessments across 452 participants. **Table 1** summarizes the demographics and subject-level average self-reported sad mood, energy, anxiousness, and sleep quality stratified by seasons. There were no statistically significant differences in the subject-level averages of the EMA variables across seasons. As expected, DMOT and daily total cloud cover significantly differed across seasons (p < 0.001), with higher total cloud cover and lower DMOT in the fall and winter.

**TABLE 1:**
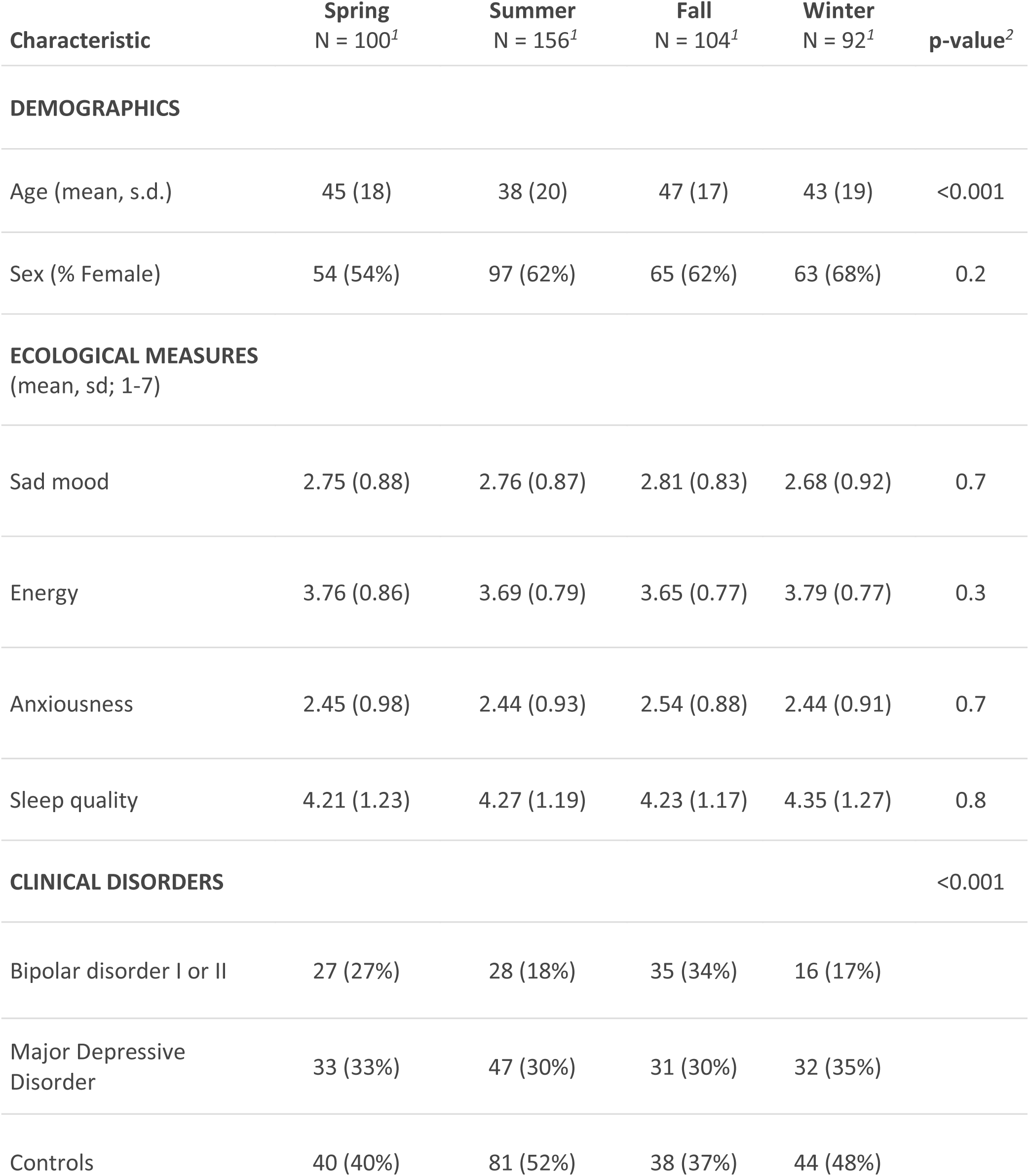
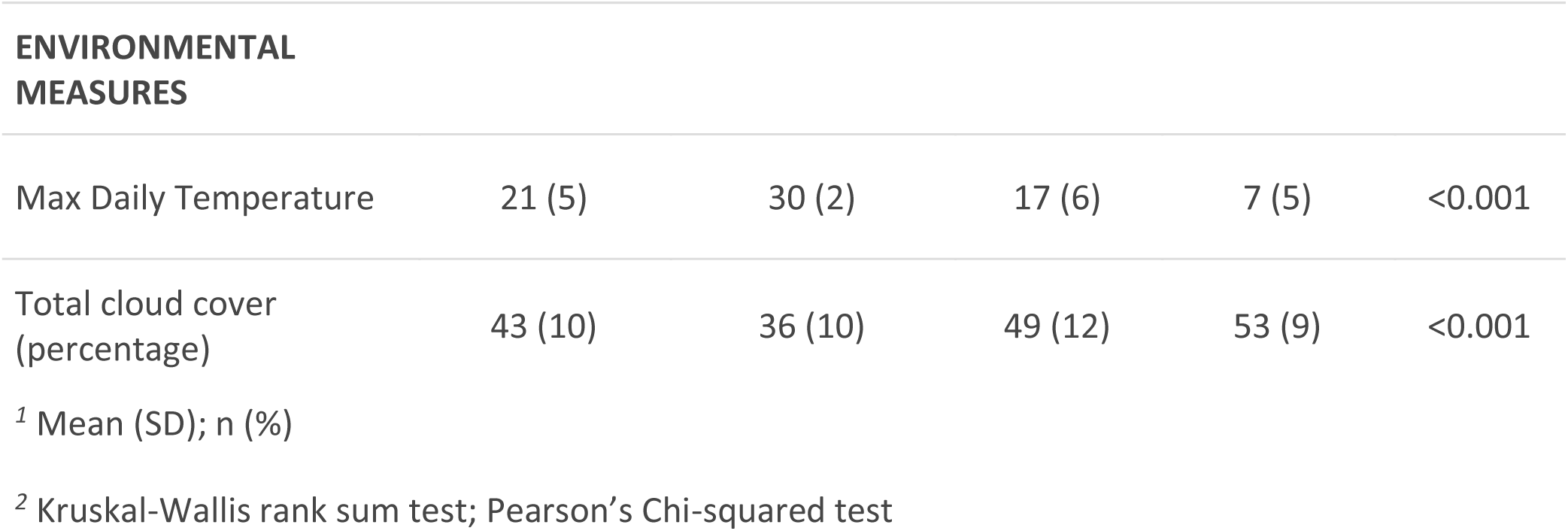
Demographics, Clinical Status, and Ecological Ratings by Season.

| Characteristic | Spring<br>N = 100<sup>1</sup> | Summer<br>N = 156<sup>1</sup> | Fall<br>N = 104<sup>1</sup> | Winter<br>N = 92<sup>1</sup> | p-value<sup>2</sup> |
| --- | --- | --- | --- | --- | --- |
| DEMOGRAPHICS |  |  |  |  |  |
| Age (mean, s.d.) | 45 (18) | 38 (20) | 47 (17) | 43 (19) | <0.001 |
| Sex (% Female) | 54 (54%) | 97 (62%) | 65 (62%) | 63 (68%) | 0.2 |
| ECOLOGICAL MEASURES<br>(mean, sd; 1-7) |  |  |  |  |  |
| Sad mood | 2.75 (0.88) | 2.76 (0.87) | 2.81 (0.83) | 2.68 (0.92) | 0.7 |
| Energy | 3.76 (0.86) | 3.69 (0.79) | 3.65 (0.77) | 3.79 (0.77) | 0.3 |
| Anxiousness | 2.45 (0.98) | 2.44 (0.93) | 2.54 (0.88) | 2.44 (0.91) | 0.7 |
| Sleep quality | 4.21 (1.23) | 4.27 (1.19) | 4.23 (1.17) | 4.35 (1.27) | 0.8 |
| CLINICAL DISORDERS |  |  |  |  | <0.001 |
| Bipolar disorder I or II | 27 (27%) | 28 (18%) | 35 (34%) | 16 (17%) |  |
| Major Depressive Disorder | 33 (33%) | 47 (30%) | 31 (30%) | 32 (35%) |  |
| Controls | 40 (40%) | 81 (52%) | 38 (37%) | 44 (48%) |  |
| ENVIRONMENTAL MEASURES |  |  |  |  |  |
| Max Daily Temperature | 21 (5) | 30 (2) | 17 (6) | 7 (5) | <0.001 |
| Total cloud cover (percentage) | 43 (10) | 36 (10) | 49 (12) | 53 (9) | <0.001 |

<sup>1</sup> Mean (SD); n (%)
<sup>2</sup> Kruskal-Wallis rank sum test; Pearson’s Chi-squared test

### Consistent associations across domains

The estimated non-linear associations between self-reported sad mood, energy, anxiousness, sleep quality, and DMOT stratified by season are presented in **Figures S1-S4**. Note that when estimating non-linear associations using penalized splines, the estimated association may be approximately linear if the data do not sufficiently support non-linearity. **Figures 1-2** summarize key significant associations in individuals with Bipolar disorder and MDD. The associations reflect the significance of the overall effects within each diagnostic group, rather than a comparison to the control group. Consistent associations were found across domains for individuals with Bipolar disorder (BD) and MDD in the fall and spring. **Table 2** summarizes findings of the DMOT associations in different EMA across different seasons and mood subgroups.

**Table 2:** Associations by Mood Subgroups, Seasons, and EMA Domains.

Season-specific effect of temperature on EMA values by Diagnostic Group
| EMA | Sad Mood |  |  |  | Energy |  |  |  | Anxiousness |  |  |  | Sleep Quality |  |  |  |
| --- | --- | --- | --- | --- | --- | --- | --- | --- | --- | --- | --- | --- | --- | --- | --- | --- |
|  | SP | SU | FA | WI | SP | SU | FA | WI | SP | SU | FA | WI | SP | SU | FA | WI |
| ALL | ↓ |  |  |  |  |  |  |  |  |  | ↑ |  | ↑ |  |  |  |
| CTL |  |  |  |  | ↓ |  | ↑ | ● | ↑ | ↑ |  |  |  |  |  |  |
| BD | ↓ |  | ● | ● | ↑ |  | ● |  |  |  | ● |  | ↑ |  | ↑ | ↑ |
| MDD | ↓ |  |  |  | ↑ |  |  | ↓ |  |  | ● |  | ↑ | ↑ |  | ↓ |

**Green Arrow (↑ or ↓):** Indicates a beneficial effect, with the direction showing the directionality of the association. **Red Arrow (↑ or ↓):** Indicates a detrimental effect, with the direction showing the directionality of the association. **Blue Circle (●):** Represents a nonlinear relationship. **Gray Cell:** Indicates no significant effect (none).

In spring, higher DMOT was associated with improved mood, increased energy, and better sleep quality in individuals with BD and MDD **(Figure 1)**. A 5-degree Celsius temperature increase from the median was associated with a 5.7% decrease in sad mood, 4.5% higher energy, and 5.4% better sleep quality in BD. In contrast, in MDD, the exact change in temperature corresponds to 4.3% lower sad mood, 2.2% higher energy, and 6.3% better sleep quality.

In fall, there was an inverse U-shaped association of DMOT with negative emotional states, such as self-reported sad mood and anxiousness, and a U-shaped association with positive emotional states, such as energy, among people with BD **(Figure 2)**. Specifically, lower and higher DMOT (compared to the seasonal median) were associated with decreased sad mood, anxiousness, and increased energy. To quantify, a 5-degree Celsius temperature increase from the median was associated with a 2.5% decrease in sad mood, 5.9% higher energy, and 2.3% less anxiousness in BD, whereas a 5-degree Celsius temperature decrease from the median was associated with a 6.7% decrease in sad mood, 5.7% higher energy, and 5.7% less anxiousness in those BD l. By contrast, there were fewer associations between temperature and mood states among those with MDD, except for the association of higher levels of anxiousness with both higher and lower DMOT.

### Domain-specific significant associations

In spring, higher DMOT was associated with reduced energy among controls **(Figure S2)**. In spring, summer, and winter, higher DMOT was associated with higher levels of anxiousness among the controls. It’s worthwhile to note that controls also include individuals diagnosed with Anxiety disorder. This effect could be partially explained only by people with Anxiety disorder, but we can’t comment further due to the low sample size.

There were fewer notable associations between temperature and emotional domains in the winter and summer than in the spring and fall (**Figures S1-S4**). People with MDD tended to be more susceptible to temperature-related emotional changes, with higher winter temperatures associated with lower energy levels (**Figure S2**). People with BD reported higher levels of sad mood with increasing DMOT in winter **(Figure S1**). In summer, higher DMOT was associated with better sleep quality in those with MDD, but in winter, higher DMOT was associated with lower sleep quality. Although there were fewer winter and summer effects in those with BD, they did exhibit better sleep quality with higher DMOT in the fall. (**Figure S4**).

## DISCUSSION

### Summary of findings and context

This work contributes to the growing evidence regarding the association of ambient temperatures with emotional states and disorders assessed in real-time across seasons (Clery et al., 2024) and provides insight into potential mechanisms for the links between temperature on serious mental health outcomes including mental health hospitalizations and deaths by suicide (Thompson et al., 2018)(Liu et al., 2021; Radua et al., 2024). In parallel to population-level statistics on hospitalization, suicide attempts, and death (Yu et al., 2020), the strongest associations with temperature emerged in spring, with significant but less pronounced effects in the fall. There were differential associations with the range of daily measures among people with a history of mood disorders, particularly those with BD, in whom temperature changes were associated with fluctuations in mood, energy, and sleep quality when compared to controls. Therefore, in this era of rapid and unpredictable changes in weather, systematic investigation of the associations between the complex elements of climate, emotions, and health is critical for understanding within-person dynamics related to temporal trends in mental and physical health.

### Differential Influence of temperature on clinical mood disorder subtypes

This work adds to the growing evidence for increased susceptibility of people with BD (Bauer et al., 2009; Clery et al., 2024; Lee et al., 2007; Shapira et al., 2004; Sung et al., 2013) to ambient temperature fluctuations, particularly more extreme variation that deviates from usual seasonal patterns (Clery et al., 2024). In the present study, the effects of temperature differed for people with a history of mood disorders in general, with greater fluctuation in mood, energy, and sleep, especially in the spring and fall. Among those with a history of a mood disorder, warmer temperatures were associated with increased mood, energy, and sleep quality in the spring. There was also a non-linear effect in the fall with both increasing and decreasing temperatures associated with lower sad mood, higher energy, and lower anxiousness in people with BD. Together, these findings suggest that the well-documented enhanced sensitivity of people with BD to daily contextual perturbations at the individual level (Merikangas et al., 2019) may extend to broader environmental conditions, in particular temperature and its correlates (Lee et al., 2007).

### Sleep and energy may explain temperature influences on mood states

These findings contribute to the growing evidence regarding aggregate environmental influences on the dynamics of mood disorders. Our inclusion of sleep quality and emotional states, such as energy beyond assessment of mood, may provide insight into the potential mechanisms for the link between temperature and mental health outcomes. In the present study, changes in sleep quality tended to co-occur with changes in mood and energy, suggesting that temperature-induced sleep disruption could explain the observed mood variation. For example, through linkage of meteorological data with mobile sleep assessments in 47,628 adults across 68 countries, Minor and colleagues (Minor et al., 2022) found that increased temperature was associated with shortened sleep primarily through delayed onset, increasing the probability of insufficient sleep, particularly among residents from lower-income countries and older adults. Recent reviews of evidence for the influence of ambient heat and sleep have shown that both higher outdoor and indoor temperatures are generally associated with degraded sleep quality and quantity worldwide (Chevance et al., 2024). In light of this evidence for the influences of temperature on both mental health in general and sleep patterns, studies of the dynamics of BD should consider both aggregate environmental conditions, in addition to individual-level triggers and predictors of episodes, in the conceptualization of risk and interventions for both physical and mental disorders.

### Limitations

This study has several limitations that should be considered when interpreting the findings. First, although we controlled for light that is highly correlated with temperature across seasons (Rudolph et al., 2019), other factors, such as humidity and ambient noise that are also associated with weather and temperature, were not included in our analyses. Second, our study is limited by our reliance on participant zip codes as a proxy of exposure to ambient temperature and cloud cover rather than direct information on individual-level outdoor exposure at the time of the EMA ratingsFuture studies that collect direct data on light, temperature, noise and humidity data through wearable detectors can enhance the accuracy of these assessments. Third, the small and unequal sample size of each specific seasonal stratum reduced the power to detect seasonal influences on these outcomes. Fourth, although our sample was community-based, it was not representative of either the general community or of clinical cases of mood disorders. Fifth, the sample was geographically restricted to the Washington, DC, Maryland, and Virginia regions, thus reducing the variability in temperature for a specific time of the year. Efforts to expand this work across the US and international sites are now underway in the Motor Activity Research Consortium for Health (mMARCH) (Guo et al., 2022), enabling us to examine these influences at different latitudes, seasons, and environmental exposures.

This work exploits the available spatial data on temperature and other aggregate environmental factors increasingly used for environmental and public health research. The influence of natural surroundings on psychological well-being is well known (Allen et al., 2014; Baylis et al., 2018). With the increasing threat of disruption of external environmental factors such as light and temperature (Berry et al., 2010; McMichael et al., 2006; Obradovich & Minor, 2022) induced by changes in weather and climate, it is paramount to understand how meteorological variables influence human mental health. Future studies of much larger samples across a broad geographic distribution (Bauer et al., 2019) and more comprehensive data on both aggregate and individual level exposures to environmental contextual factors will be valuable in gaining insight into mechanistic explanations that may enable the identification of individuals with the greatest vulnerability to climate change on physical and mental health.

## Ethical standards

Research was conducted under clinical protocol 03-M-0211 (NCT00071786). All participants signed informed consent.

## AUTHOR CONTRIBUTIONS (CRediT)

Conceptualization: DD, KRM; Data curation: DD, HAL; Formal analysis: DD, AL, VZ; Funding acquisition: KRM; Investigation: DD, KRM; Methodology: DD, AL, VZ; Project administration: KRM; Resources: KRM; Software: DD, AL; Supervision: KRM; Validation: DD, AL; Visualization: DD; Writing – original draft: DD; Writing – review & editing: all authors.

## Data Availability

All data produced in the present study are available upon reasonable request to the authors

## Acknowledgments

None

## SUPPLEMENT

### Section 1: Figures

**Figure S1:**
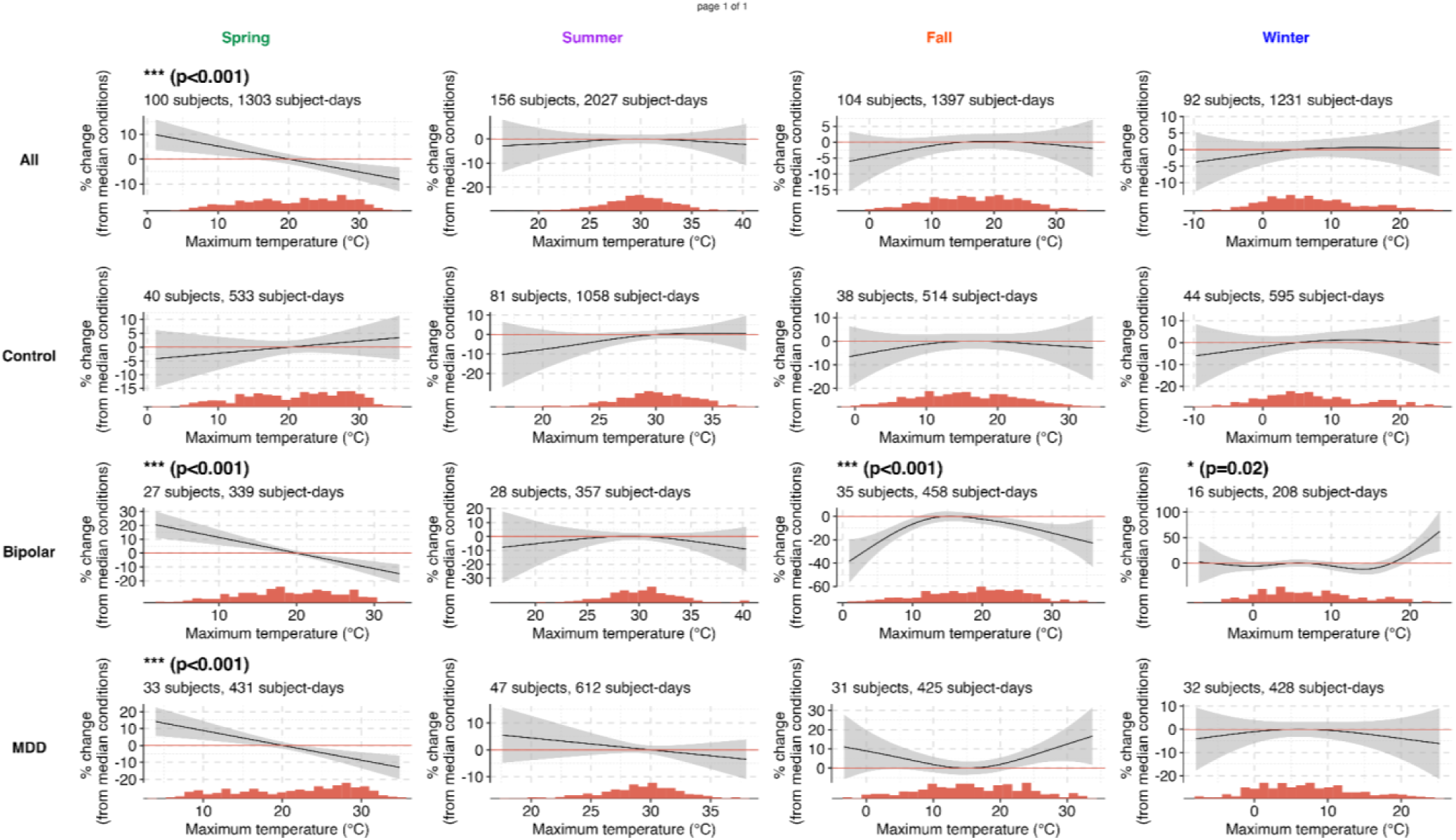
Percentage change in Sad Mood with DMOT stratified across seasons and history of mood disorder subtypes.

**Figure S2:**
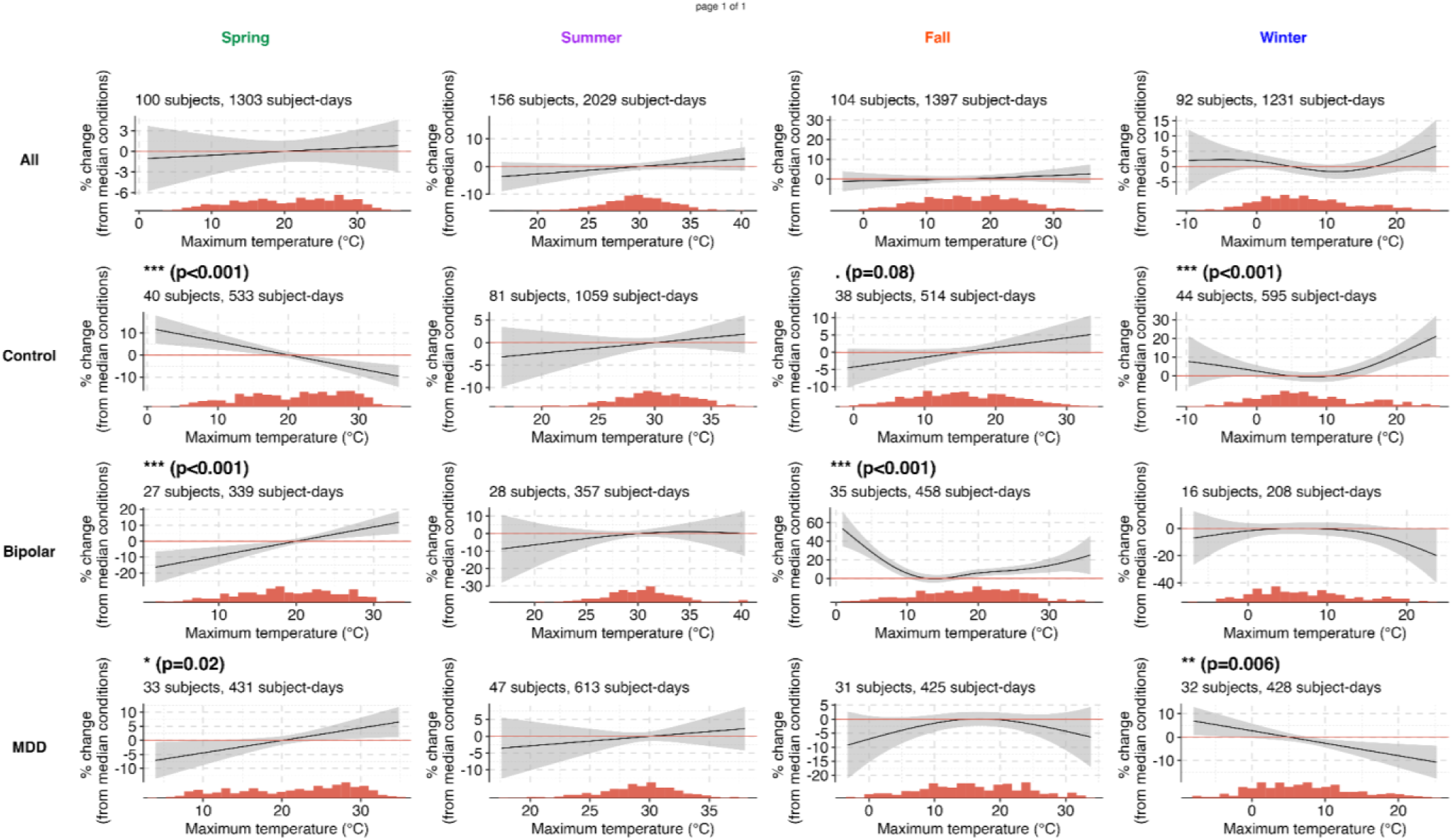
Percentage change in ENERGY with DMOT stratified across seasons and history of mood disorder subtypes.

**Figure S3:**
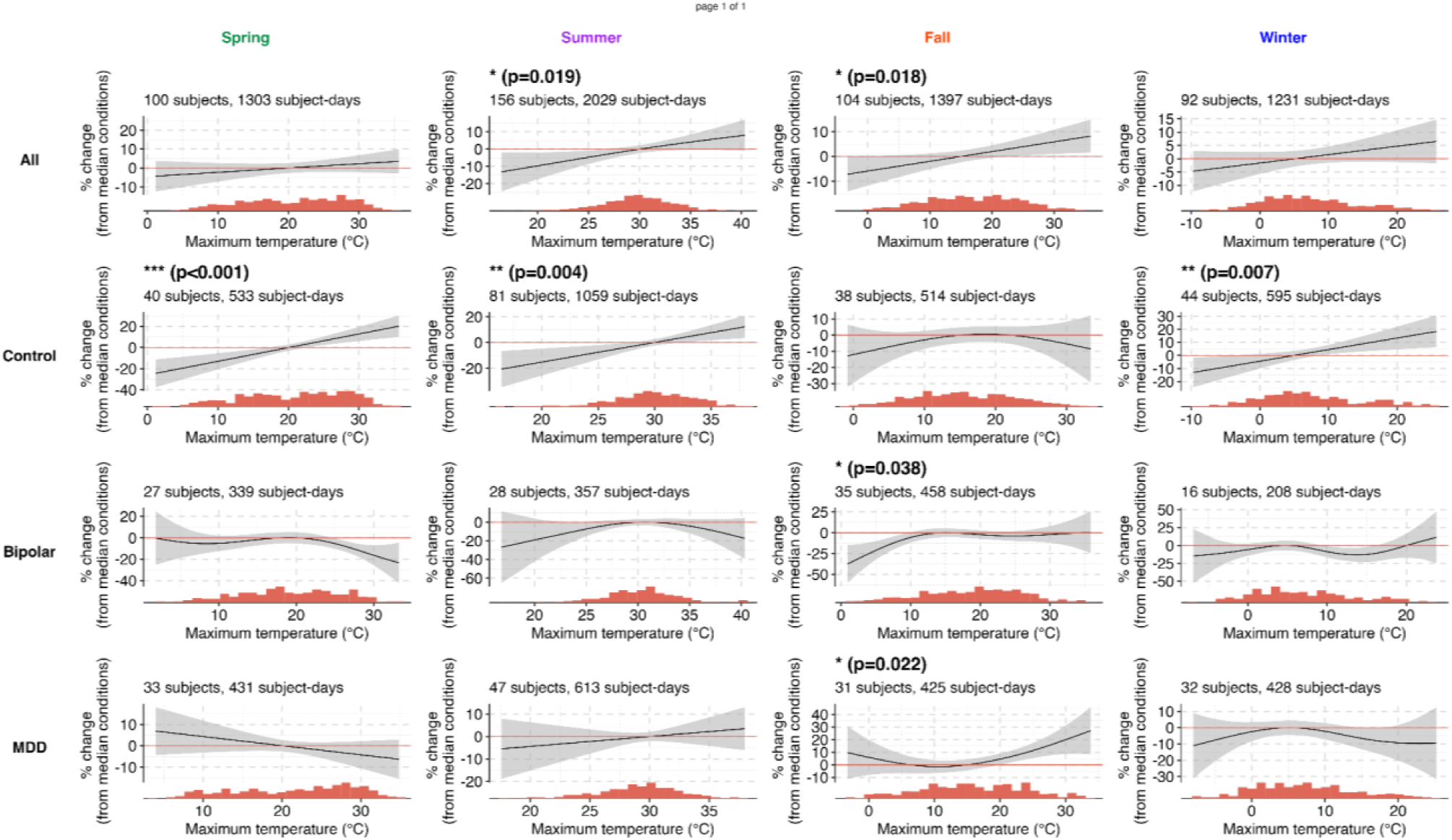
Percentage change in ANXIOUSNESS with DMOT stratified across seasons and history of mood disorder subtypes.

**Figure S4:**
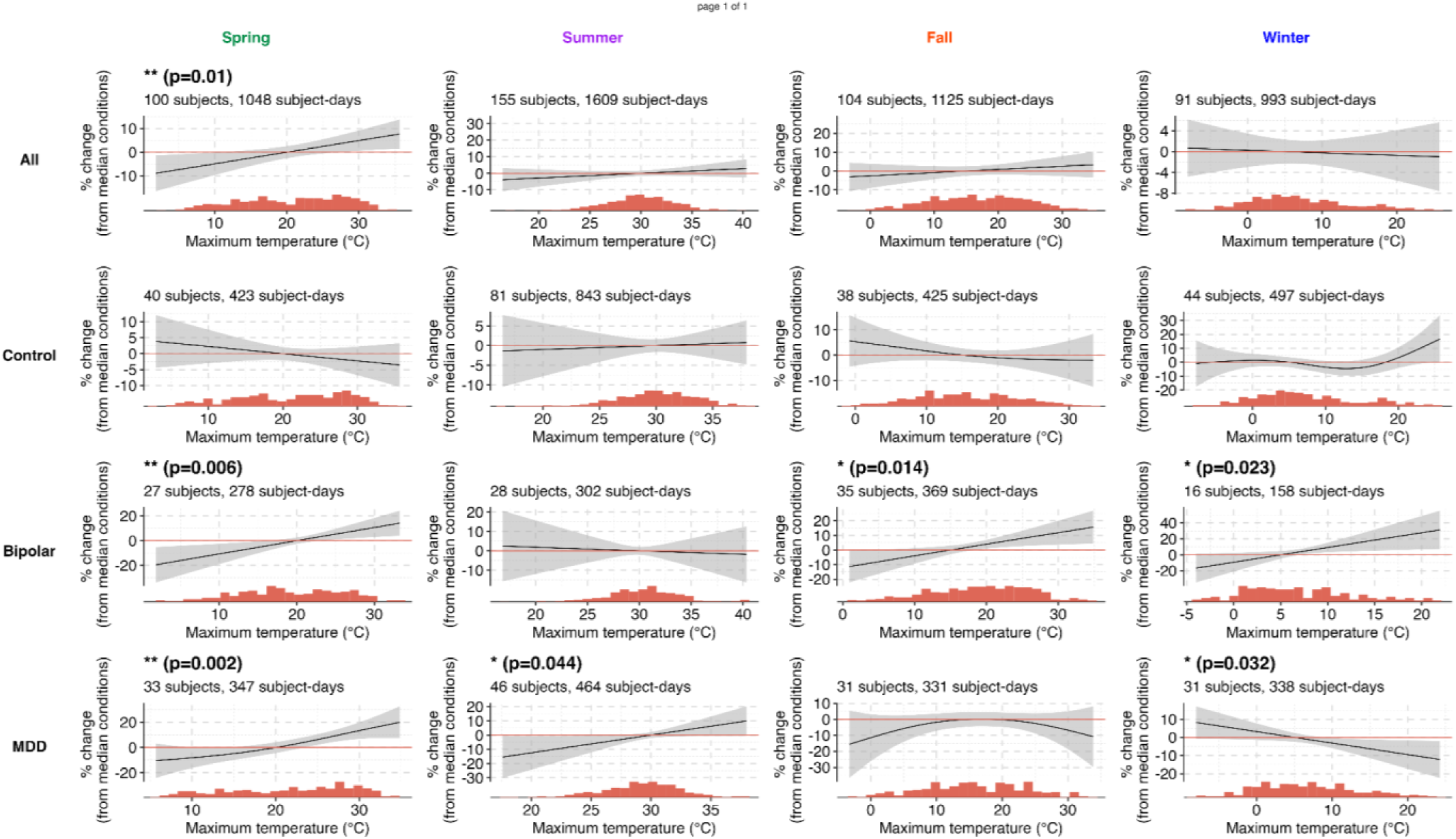
Percentage change in SLEEP QUALITY with DMOT stratified by seasons and the history of mood disorder subtypes.

### SECTION 2: STATISTICAL MODELS

**Model set *M***^(***s***)^, s = {spring, summer, fall, winter}:

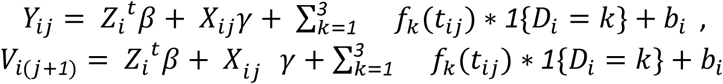

Model set ***M***^(***s***)^denotes the set of models fit on the subsample of subjects from a specific season *s*. Here, *Y*_*ij*_ is the average daily value for an EMA variable (self-reported sad mood/ energy/anxiousness) on day *j* for participant *i* and *V_i(j+1)_* is the self-reported sleep quality for (j + *1*)-th day for participants *i*. *Z*_i_ and *β* are the design matrix and regression coefficients associated with non-time-varying adjusters (sex, age, history of mood disorder, and an indicator for whether day *j* is a weekend (Friday/Saturday/Sunday)), *b*_i_ is a subject-specific random intercept which accounts for within-subject correlation, *X*_*ij*_ is the average daily total cloud cover (in percentage) and *t*_*ij*_ is the daily maximum outdoor temperature (DMOT) on day *j* for participant *i*. *f*_*k*_’s denote smooth functions of temperature modeled as a penalized B-spline. *D*_i_ denotes the history of mood disorder for the participant i - where *D*_i_=1 for the **Control** group, *D*_i_=2 for the **Bipolar** group, and *D*_i_=3 for the **MDD** group. The models are fitted using the MGCV package (S. Wood & Wood, 2015) in R Statistical software.

